# Monitoring and forecasting the number of reported and unreported cases of the COVID-19 epidemic in Brazil using Particle Filter

**DOI:** 10.1101/2020.05.27.20115212

**Authors:** J. C. S. Dutra, W. B. da Silva, J. M. J. da Costa

**Affiliations:** Chemical Engineering Program, UFES, Alegre, 29500-000, Brazil; Statistics Department, UFAM, Manaus, AM, CEP 69077-000, Brazil

**Keywords:** Epidemics modelling, SIRU model, COVID-19, Particle filter, Particle swarm optimization, Bayesian inference

## Abstract

In this paper, we combine algorithm of Liu & West for the Particle Filter (PF) with SIRU-type epidemic model to monitor and forecast cases of Covid-19 in Brazil from February up to September. We filter the number of cumulative reported cases and estimate model parameters and more importantly unreported infectious cases (asymptomatic and symptomatic infectious individuals). The parameters under study are related to the attenuation factor of the transmission rate and the fraction of asymptomatic infectious becoming reported as symptomatic infectious. Initially, the problem is analysed through Particle Swarm Optimization (PSO) based simulations to provide initial guesses, which are then refined by means of PF simulations. Subsequently, two additional steps are performed to verify the capability of the adjusted model to predict and forecast new cases. According to the results, the pandemic peak is expected to take place in mid-June 2020 with about 25,000 news cases per day. As medical and hospital resources are limited, this result shows that public health interventions are essential and should not be relaxed prematurely, so that the coronavirus pandemic is controlled and conditions are available for the treatment of the most severe cases.

## 1 Introduction

The first infection by the new coronavirus (Sars-CoV-2) was detected on December 31, 2019, in Wuhan, China and a pandemic was declared by the World Health Organization on March 11, 2020 [1, 2]. Such infection causes the Covid-19, whose main symptoms include cough, fever, difficulty breathing. It presented a scary evolution with millions of infected people and thousands of deaths in the world. Because these facts, the affected countries and cities around the world have been reacting in different ways, towards locally controlling the disease evolution.

In Brazil, the first case of Covid-19 was confirmed on February 26, 2020. Ever since, the number has surged to more than 310,000 cases of the disease, according to the Brazilian Ministry of Health [3]. Such milestone makes this nation to become the third one with the highest number of infections in the world, behind only the United States and Russia. Specifically, Amazonas, Ceara, Pernambuco, Rio de Janeiro and Sao Paulo have been the worst affected states in Brazil, whose cases correspond to at least 60% of all cases in the country. The coronavirus has so far been responsible for more than 20,000 deaths. On May 21, 2020, an unenviable record was broken with 1,188 new deaths reported officially within a single day. This acceleration of the pandemic shows that Brazil tends to become a new global epicentre of the covid-19 [4].

Figure 1 comprising data up to May 21, 2020 shows that the number of confirmed cases of the Covid-19 in Brazil has significantly exceeded the number of cases in China. This extent of the disease reveals a dramatic situation, since the population of Brazil is approximately 6.5 times smaller than that of China, which implies the urgent need for epidemiological studies to develop the best strategies for public health policies.

**Figure 1:**
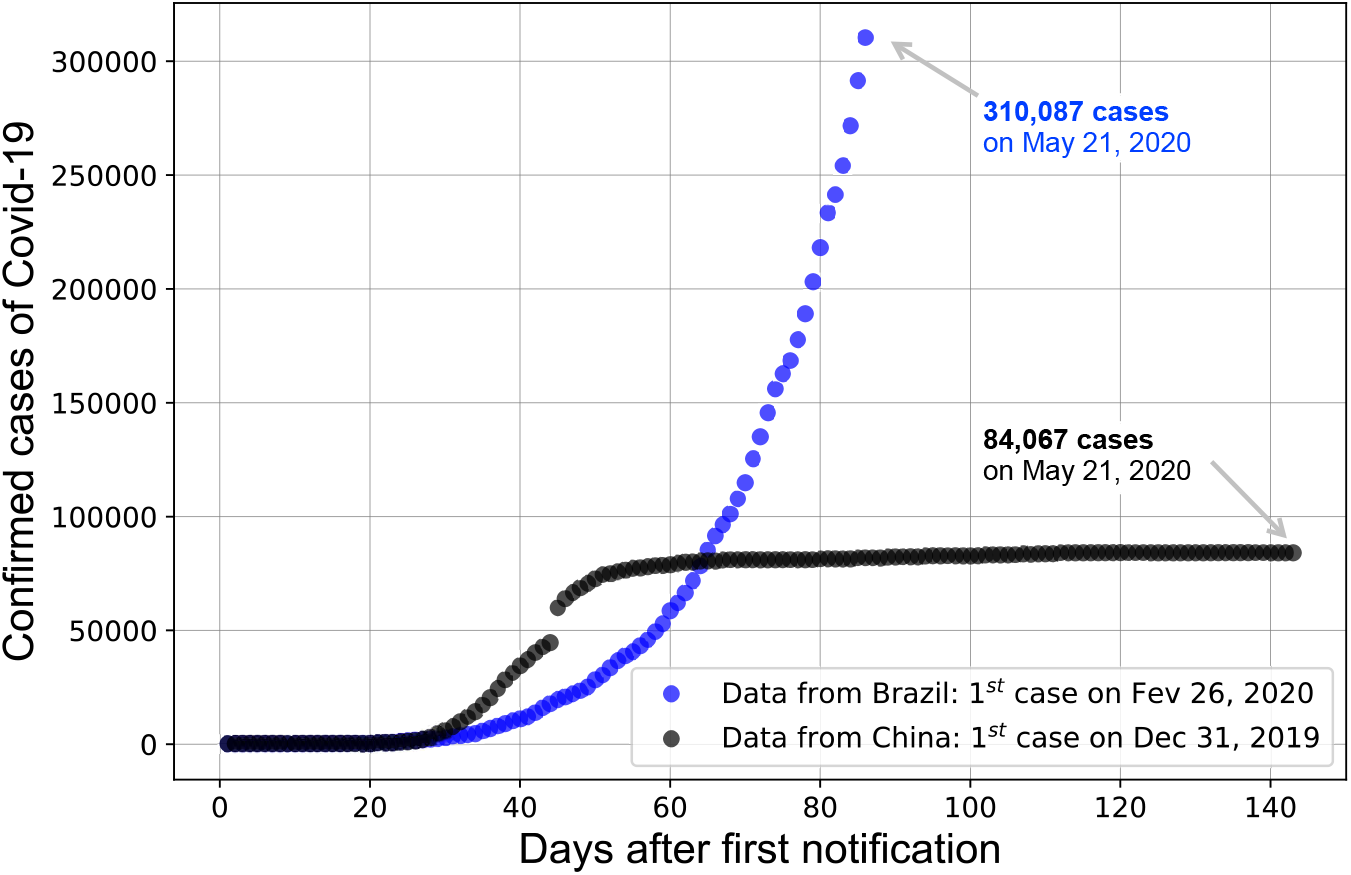
Comparison between the number of confirmed cases of Covid-19 in Brazil (85 days since first case) and in China (142 days since first case) up to May 21, 2020.

Public strategies include general isolation through quarantine and massive testing for focused isolation, with varying degrees of success so far, as can be analysed from the limited data available [5]. In this regard, mathematical modelling is an interesting approach that can allow the evaluation of different scenarios, furnishing information for a proper support for health system decisions. In general, nonlinear dynamics of biological and biomedical systems is the objective of several researches that can be based on mathematical modelling or time series analysis [6]. Literature presents some examples related to the dynamics of infectious diseases. Mathematical equations are widely used to model the nature and impact of global pandemics in the society. In particular, coronavirus propagation can be described by a mathematical model that allows the nonlinear dynamics analysis, representing different populations related to the phenomenon [7]. Recently, the classical susceptible-infectious-recovered (SIR) model proposed in [8, 9], was employed in the analysis of the epidemic outbreak in different countries, including China, South Korea, Germany, Italy, and France [10, 11, 12]. The modelling and evaluating the consequences of public health interventions. It was a direct application of previous developments [13, 14] on the fundamental problem of parameter identification in mathematical epidemic models, accounting for unreported cases.

In this work the SIRU-type model is implemented for the direct problem formulation of the COVID-19 epidemic evolution in Brazil, adding a time variable parametrization for the fraction of asymptomatic infectious that become reported symptomatic individuals. According Cotta *et al*. [5] it is an especially important parameter in the public health measure associated with massive testing and consequent focused isolation. The same analytical identification procedure is maintained for the required initial conditions, as obtained from the early stage exponential behaviour. However, a Bayesian inference approach is here adopted for parametric estimation, employing the combined parameter and state estimation algorithm proposed by Liu and Wes and Particle Swarm Optimization (PSO).

## 2 Mathematical formulation

In this section, we formulate the mathematical SIR model [5, 9, 10, 11, 12, 13, 14] we analyze is the following: at time *t*, let *S*(*t*) be the number of susceptible, *I*(*t*) the number of infected, *R*(*t*) the number of removed and permanently immune, *U* (*t*) and number of unreported symptomatic infectious individuals. The equations for the SIR model

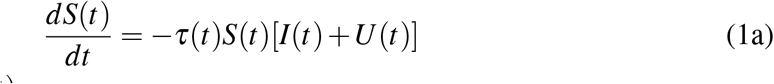

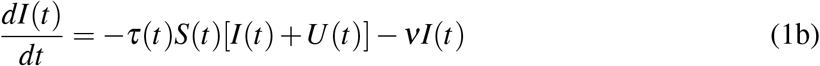

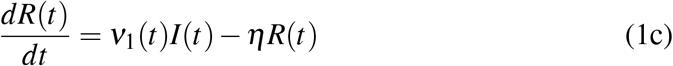

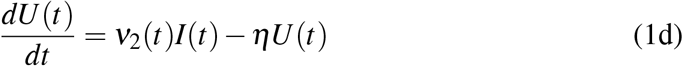

where,

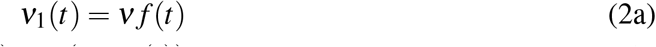

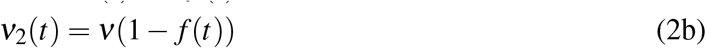

with initial conditions

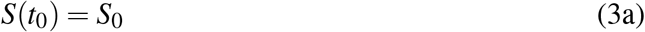

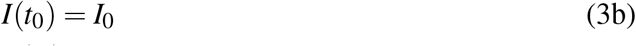

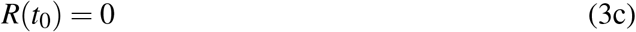

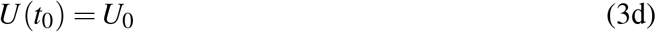

− where *t*_0_ is the beginning date of the epidemic in days. Asymptomatic infectious individuals *I*(*t*) are infectious for an average period of 1*/ν* days. Reported symptomatic individuals *R*(*t*) are infectious for an average period of 1*/η* days, as are unreported symptomatic individuals *U* (*t*). According to Cotta *et al*. [5] assumed that reported symptomatic infectious individuals *R*(*t*) are reported and isolated immediately, and cause no further infections. The asymptomatic individuals *I*(*t*) can also be viewed as having a low-level symptomatic state. All infections are acquired from either *I*(*t*) or *U* (*t*) individuals. The fraction *f* (*t*) of asymptomatic infectious become reported symptomatic infectious, and the fraction 1 − *f* (*t*) become unreported symptomatic infectious. The rate asymptomatic infectious become reported symptomatic is *ν*1(*t*) = *f* (*t*)*ν*, the rate asymptomatic infectious become unreported symptomatic is *ν*_2_(*t*) = (1 − *f* (*t*)), where *ν*_1_(*t*) + *ν*_2_(*t*) = *ν*. The transmission rate, *τ*(*t*), is also allowed to be a time variable function along the evolution process. Figure 2 below illustrates the infection process as a flow chart [10].

**Figure 2:**
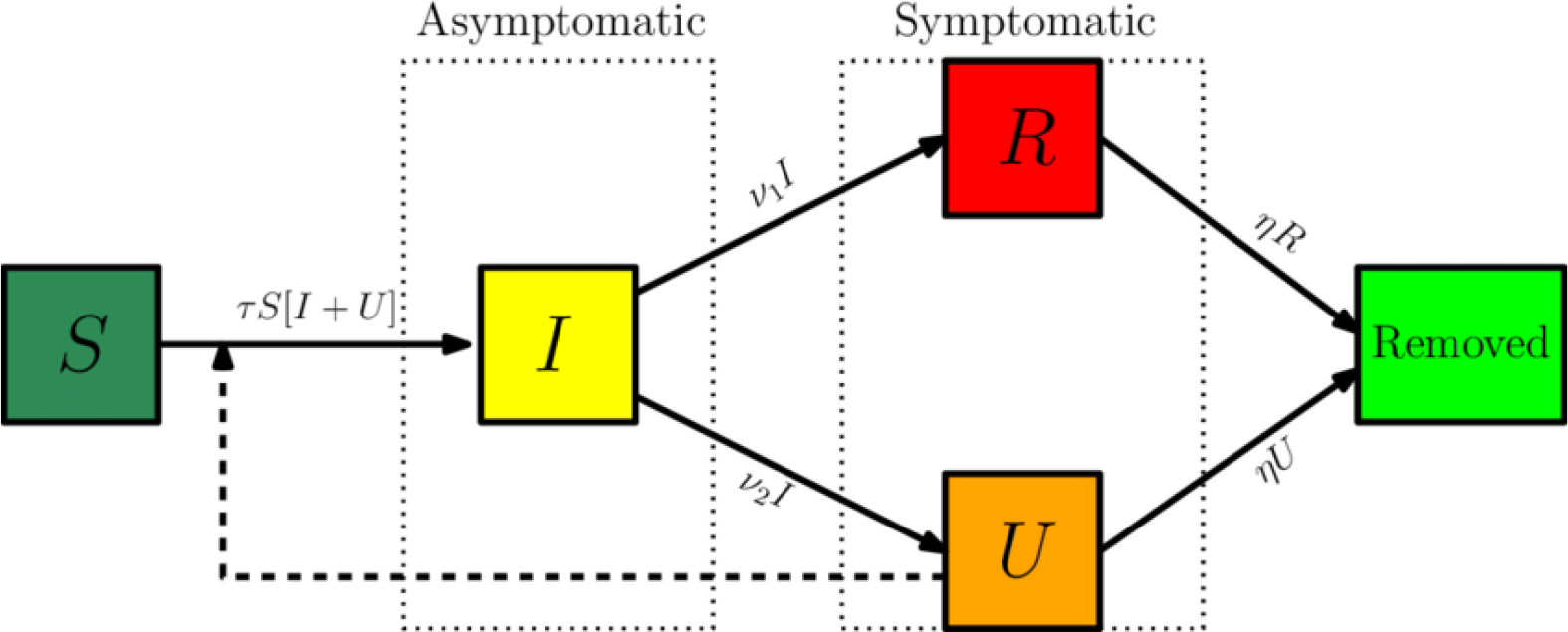
Flow chart illustrating the infection path process.

The time variable coefficients, *τ*(*t*) and *f* (*t*), are chosen to be expressed as:

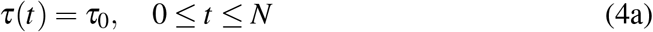

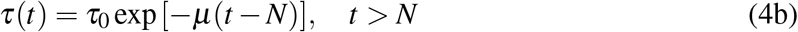

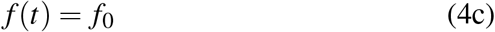

These parameterized functions are particularly useful in interpreting the effects of public health interventions. For instance, the transmission rate, *τ*(*t*), is particularly affected by a reduced circulation achieved through a general isolation or quarantine measure, while the fraction *f* (*t*) of asymptomatic infectious that become reported, thus isolated, cases can be drastically increased by a massive testing measure with focused isolation. In the above relations, *µ* is the attenuation factor for the transmission rate, *N* is the time in days for application of the public health intervention to change transmission rate, *µ_f_* is the argument of the *f* (*t*) variation between the limits(*f*_0_, *f_max_*) e first time variable function has been previously considered, while the second one has been introduced in the present work, so as to allow for the examination of combined measures.

The cumulative number of reported cases at time *t, CR*(*t*), which is the quantity offered by the actual available data, and the a priori unknown cumulative number of unreported cases, *CU* (*t*), are given by:

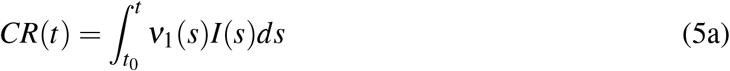

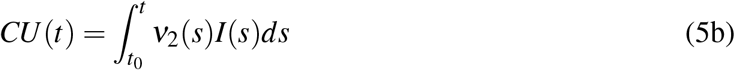

The daily number of reported cases from the model, *DR*(*t*), can be obtained by computing the solution of the following equation and with initial conditions:

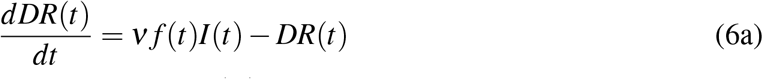

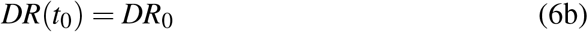

## 3 Estimation problem

The estimation problem, also known inverse problems, is an interdisciplinary task involving experimental observations and numerical analysis in many fields of science and engineering. Specifically in epidemiological studies, a crucial step is the determination of the extension of diseases. In this regard, the effort is made to infer the values of unknown states and parameters characterizing the system under investigation. The results of such studies can support developing public health policy. Here, using the available data of infected individuals in Brazil, we aim at solving a joint estimation of states and parameters (namely *τ*0, *f*0 and *µ* for the SIRU model) in order to assess the COVID-19 spread and, hence, to be able to predict locally the future progression of the pandemic.

The estimation problem, also known as non-stationary inverse problems usually requires a system dynamic model, which provides the temporal evolution of the states and parameters, an observation model, which transforms the system variables to give the observed variables, and a estimation technique that relates the all available information to the process knowledge to obtain the desired estimate [15].

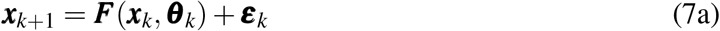

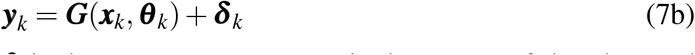

Here, ***x*** represents the state vector, ***θ*** is the parameter vector, ***y*** is the vector of the observed variables, ***ε*** and ***δ*** are respectively the system and observation noise vectors. The function ***F*** indicates the evolution model and the function ***G*** is a observation model, both of them are non-linear function of state variables and parameters. It is assumed that the noise vectors have zero mean and follow a normal distribution. The subscript *k* denotes the instant of time.

There are many numerical methods that can approach the state and parameter estimation. However, as the model considered here is nonlinear and there may be high correlation between model parameters, local optimization methods such as gradient and direct search algorithms are not suitable since they strongly depend on the initial conditions. It is necessary to resort to the use of global methods capable of handling nonlinear systems, such as:

1. Heuristic methods, including the Particle Swarm Optimization (PSO) technique which usually presents better performance than other heuristic algorithms [16, 17]; and
2. Sequential Monte Carlo methods, as the Particle Filter (PF) algorithms which makes use of random samples for a Bayesian statistical inference.

Such methods do not suffer from instability as they do not solve the inverse problem as an optimization problem, but as a search or sampling problem [18]. In both classes of methods, a set of particles is used to find the solution for the estimation problem, i.e filtering the available data and estimating the parameter values that minimize the objective function, *Fob j*, which represents the model performance for the problem. In this work, we solve the estimation problem for the SIRU model considering the measurements as the cumulative number of reported cases (*CR*(*t*)) from Brazil, using PSO and PF algorithms presented below.

Before carrying on the estimation problem, useful information can be obtained if the onset of the pandemic is modelled as an exponential growth, as represented by the following equation

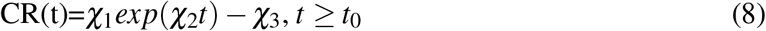

After determining *χ*_1_, *χ*_2_ and *χ*_3_, it is possible to explicitly calculate the unknown initial conditions for the model variables, estimates for the initial time of the pandemics and for the initial transmission rate [10, 9]:

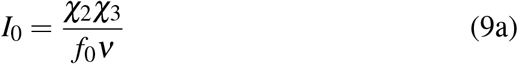

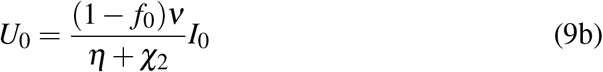

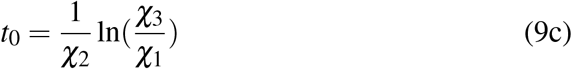

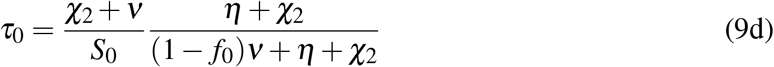

### 3.1 Particle Swarm Optimization algorithm

This technique was proposed by Kennedy and Eberhart [17] and improved by other researchers to assure the convergence to the global optimal solution. It considers that each individual of the swarm, called particle, representing a potential solution moves around in the multidimensional search space. Balancing individuality and sociability of the particles, in order to locate the optimal solution, PSO mimics the social behavior of various species adjusting dynamically the position 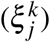 and velocity 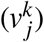 for each particle *j* = 1*,…, M* at each iteration *k*, in accordance with the following equations:

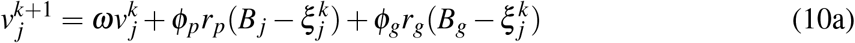

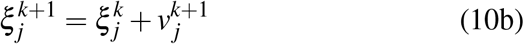

In this formulation, *j* denotes the particle, *k* represents the iteration number, *v* is the velocity and *ξ* is the position of the particle. Regarding the regions of the search space where the objective function *F_obj_*(*ξ_j_*) shows optimum values, *B_j_* is the best value found by the particle itself and *B_g_* is the best value found by the entire swarm. The coefficients *r_p_* and *r_g_* are uniform random numbers between 0 and 1; 0 < *ω* < 1 is related to inertial weight; and finally *φ_p_* and *φ_g_* denote respectively individual cognition and the social parameters that must be chosen.

A general description of the PSO algorithm follows [17, 16]:

#### Algorithm 1: The Particle Swarm Optimization.

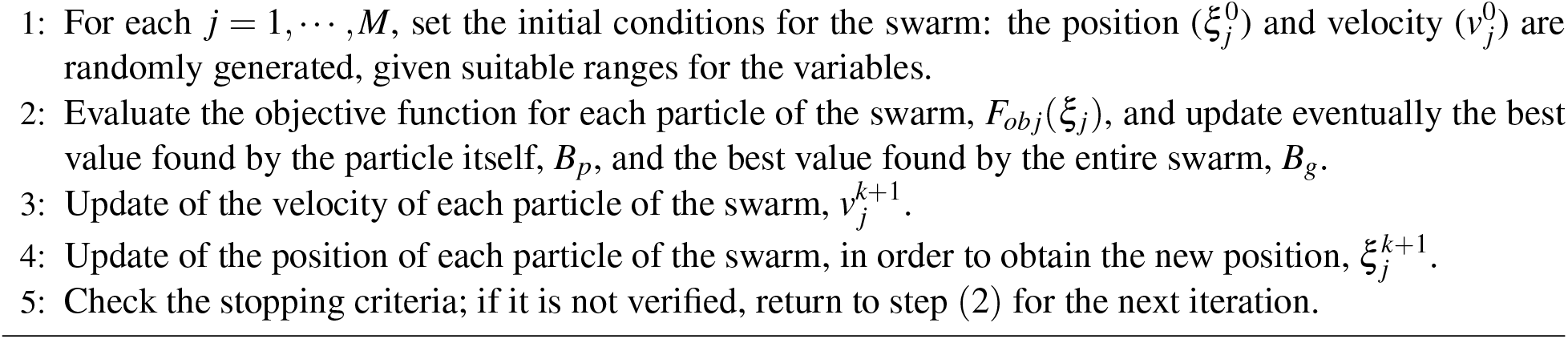

### 3.2 Particle Filter algorithm

The estimation problem with PF methods is tackled in the form of a stochastic process, in which a set particles (or samples) with associated weights is to approximate the posterior density of the states and parameters. Within the Bayesian framework, all available information (measurements and phenomena involved) is used as a source of knowledge to determine the unknown variables. The prior distribution provides the necessary information to draw the first set of particles, which are updated through the evolution model and transformed by the observation model to give the estimates. The likelihood function is then used to compare such estimates with the experimental measurements, and incorporates more information, via particle weights, in order to determine the posterior distribution. Sequentially, all new information is combined with the previous information to create the basis for the statistical procedure. Such steps describe briefly the Sequential Importance Sampling (SIS) filter [19, 15, 20, 21].

The performance of the SIS filter may be negatively affected by the degeneracy phenomenon, in which few particles contribute to the approximation of the posterior probability distribution [19, 20]. In this regard, the Sampling Importance Resampling (SIR) and Auxiliary Sampling Importance Resampling (ASIR) filters consider a resampling step to deal with the elimination of particles originally with low weights and the replication of particles with high weights, at each instant *tk* or whenever the number of effective particles falls below a certain threshold [22, 23, 24, 25, 26]. However, such filters are used to obtain estimates of state variables [19, 20].

In this work, we are particularly interested in the joint estimation of parameters and state variables, since estimating the parameters that regulate the dynamics of growth in the number of infected individuals is also very important to understand the phenomenon. For this purpose, we used the algorithm proposed by Liu and West [27]. This algorithm is a generalization of the ASIR filter, and the inference is made on the joint posterior density *π*(***x**_k_, **θ D**_k_*),where ***θ*** is the vector of parameters, ***D**_k_* contains the measurements up to time *k* and ***X**_k_* the state variables. Time evolves to *k* + 1, we observe ***y***_*k*+1_ and now want to generate a sample from *P*(***x***_*k*+1_, **θ *D***_*k*+1_). Bayes’ theorem gives this as

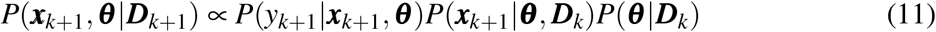

The filter is based on the hypothesis of [28], which assumes that, for a vector of static parameters ***θ***, the posterior density estimation *P*(***θ*** |***D***_*k*_) is done by smoothed density via kernel, [27]

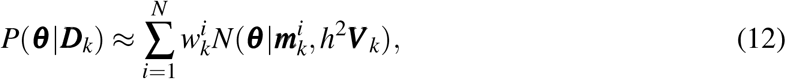

thus the artificial distribution of the parameters ***θ*** is represented by the 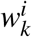 weighted mixture of normal distributions with mean ***m**_k_* and variance ***V**_k_*, where *N*(°|***m, S***) is a multivariate normal density mean ***m*** and variance matrix ***S***, where,*h* is a smoothing parameter, be chosen as a smooth and decreasing function of *N*, e

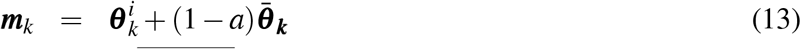

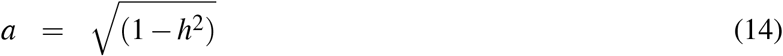

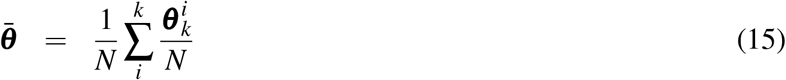

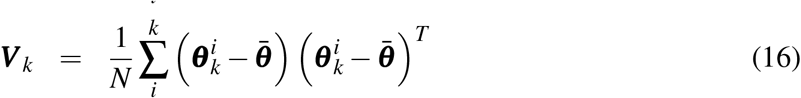

where *T* denotes the transpose of the matrix, 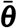 is the mean of the posterior particles. In addition, *a* is related to a discount factor *δ* as follows,[27]

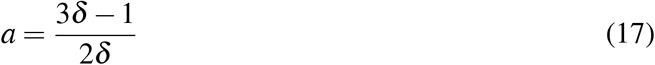

where 0.95 *< δ* < 0.99

The general particle filter algorithm for combined estimation of parameters and state variables in time step *k* − 1, based on the measurements available in time *k* is given below, [27]

## 4 Numerical results and discussion

In all simulations presented here, we assumed the estimates 1*/ν* = 6.20798 and 1*/η* = 11.2784 days [5] for the average times in the model, and also set *S*0 = 210.15 106 as the total of susceptible individuals in Brazil. Firstly, we used the data points related to the first month of the cumulative cases in Brazil, from February, 26 2020 up to March 25, 2020 (30 days), to fit the early exponential growth. In the sequence, we estimated the model parameters via inverse problem analysis applying data from February 26, 2020 up to May 21, 2020 (85 days).

Besides that, we also considered that isolation measures were taken from March 17, 2020, when domestic isolation, remote work and shutting down temporarily businesses and services were recommended. The data points recorded from May 8, 2020 until May 21, 2020 (14 days), were used to verify the predictive capability of the proposed approach. This way, we used about 84% of the data set to estimate the SIRU model parameters (learning step) and 16% for validation, changing the estimated values, if necessary, of *f*0 or *µ* to improve the model performance (prediction step). Such computational studies for direct and inverse problems were performed in PYTHON on a computer with Intel Core i5 processor and 8 GB RAM.

### 4.1 Early exponential growth model

The adjustment for the early growth of the cumulative infectious cases in Brazil is displayed in Figure 3. We can see a satisfactory agreement between the data and the model, making clear that this phase of the pandemic is purely exponential. The estimated parameters for the model *χ*_1_ = 4.185, *χ*_2_ = 0.231, and *χ*_3_ = 1.0. Such result allowed obtaining an initial time for the growth at *t*_0_ = −6.203. According to this model, a negative value for *t*0 means that the starting time of the epidemic in Brazil took place about six days before the first reported case, what may have happened on February 20, 2020. Although such modelling cannot represent subsequent cases, we can use its parameters to find the remaining initial conditions, *I*_0_, *U*_0_ and *τ*_0_, which have to be recalculated from within both PSO and PF algorithms.

#### Algorithm 2: The Liu and West Algorithm.

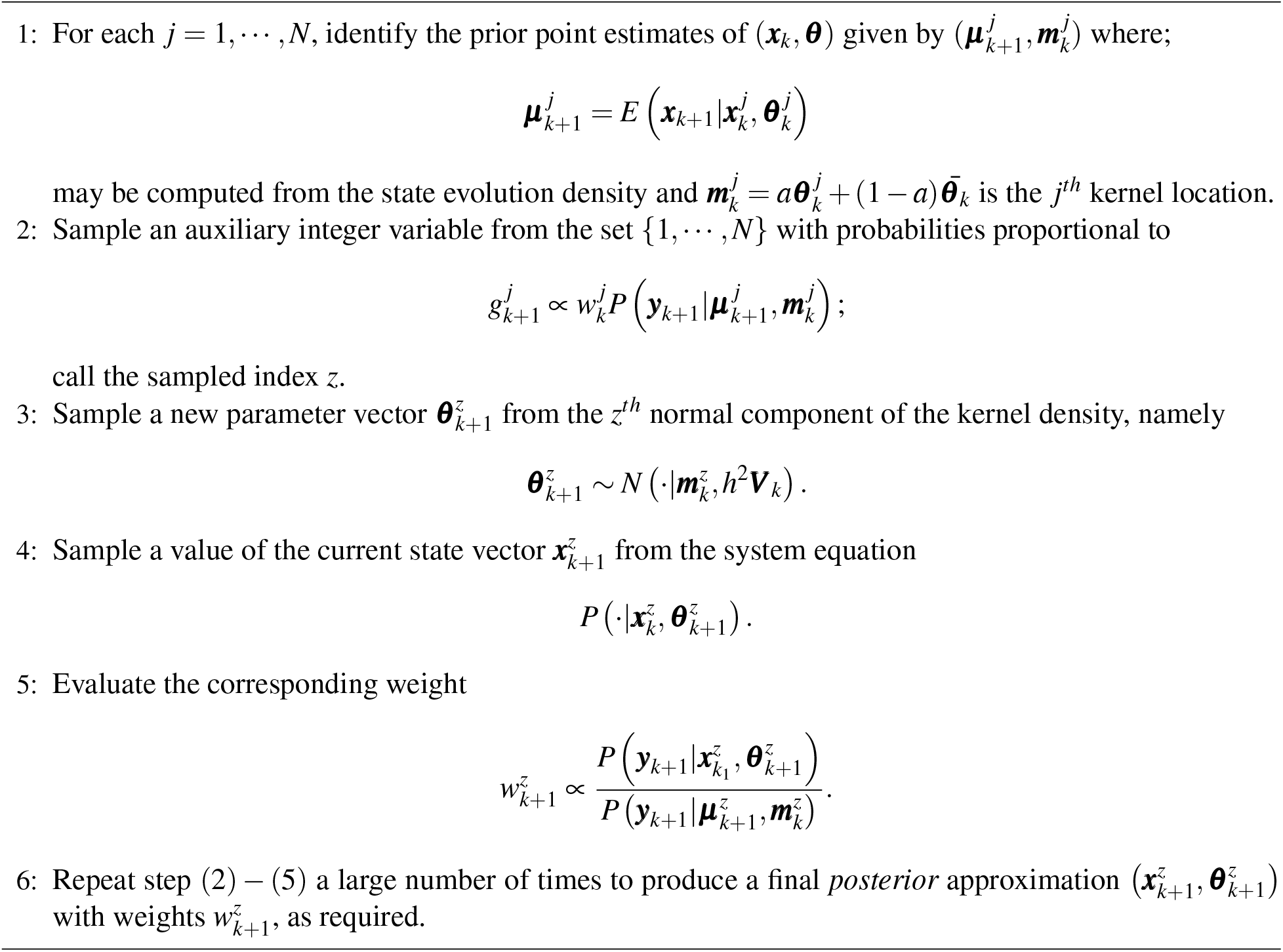

**Figure 3:**
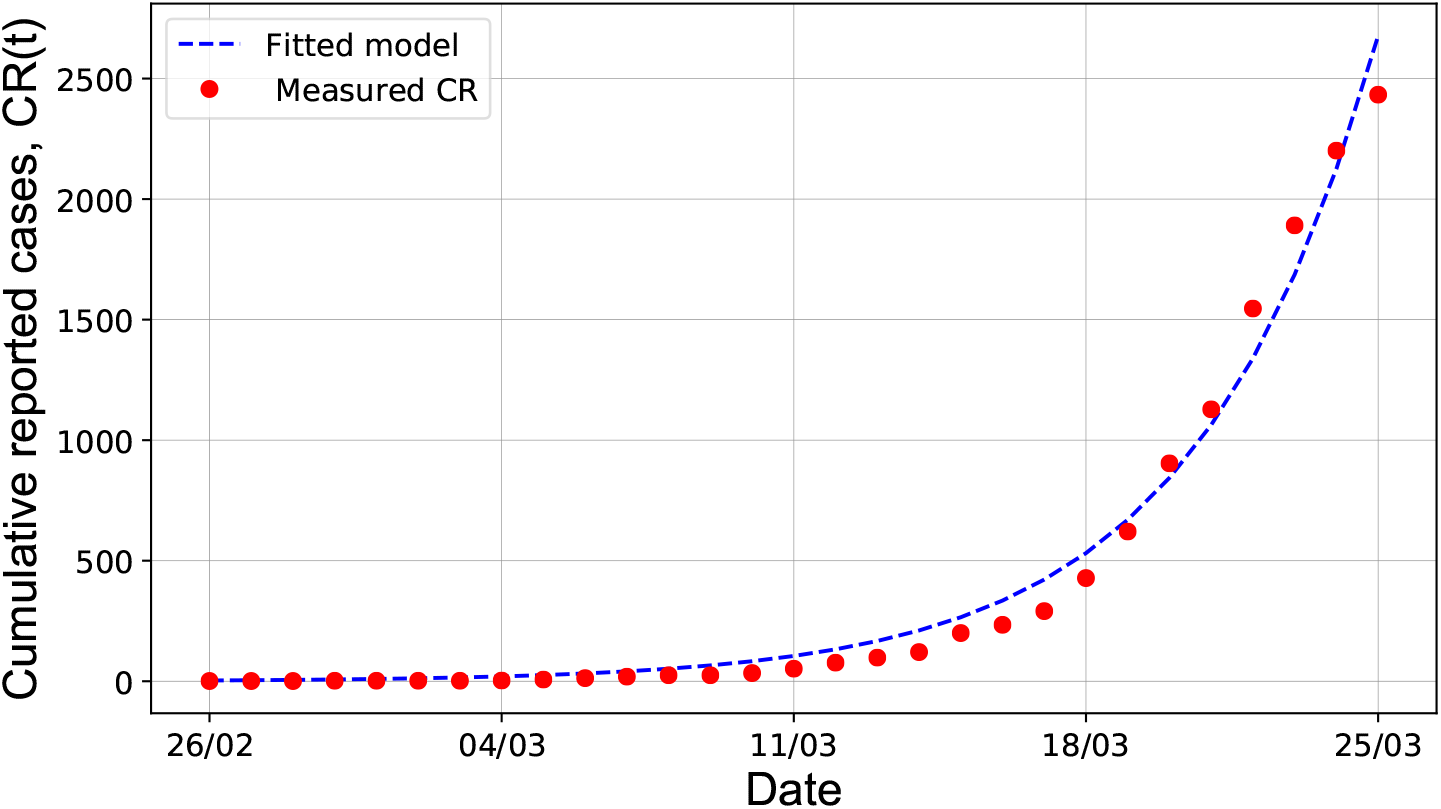
Early stage fitting for the cumulative reported cases, *CR*(*t*) = *χ*_1_*exp*(*χ*_2_*t*) − *χ*_3_, in Brazil from February, 26 2020 up to March 25, 2020, with *χ*_1_ = 4.185, *χ*_2_ = 0.231, and *χ*_3_ = 1.0.

### 4.2 Estimation from PSO simulations

To solve the estimation problem through the PSO technique, we applied a time weighted cost function as the objective function *F_obj_* = ∫ *t* ° (*CR_meas_ − CR_model_*)^2^*dt*, where *CR_meas_* and *CR_model_* are the values of the measured and estimated cumulative cases at time *t*. We used 100 particles and 50 independent calls of the code. To tune the algorithm, we set the parameters *ω* = 0.9 and *φ_p_* = *φ_g_* = 2.0[16].

Firstly, to monitor the number of reported and unreported cases and infectious cases, we used the full data set (85 days) for estimation of the SIRU model parameters, *f*_0_, *µ* and *τ*(*t*) = *g*(*t, τ*_0_*, µ*). The adjustment with PSO simulations led to the parameters *f*0 = 0.39123, *µ* = 0.0109 and *τ*_0_ = 1.426 ° 10^−9^. In Figure 4, it is possible to note an excellent agreement of the simulated values with the measured values of the cumulative reported cases (Figure 4-a). However, the number of reported cases is not represented accurately, what can also be better observed in the Figure 4-b for the daily cases. In addition, the low value of the fraction *f*0 indicates that about 40% of asymptomatic individuals became reported symptomatic infectious, meaning that a vast majority of the asymptomatic population was not diagnosed. However, the significant under reporting of cases are caused by low number of tests being carried out, delay in the official processing of information and more seriously the lack of test kits. Such reasons make it difficult to fight Covid-19 in Brazil.

**Figure 4:**
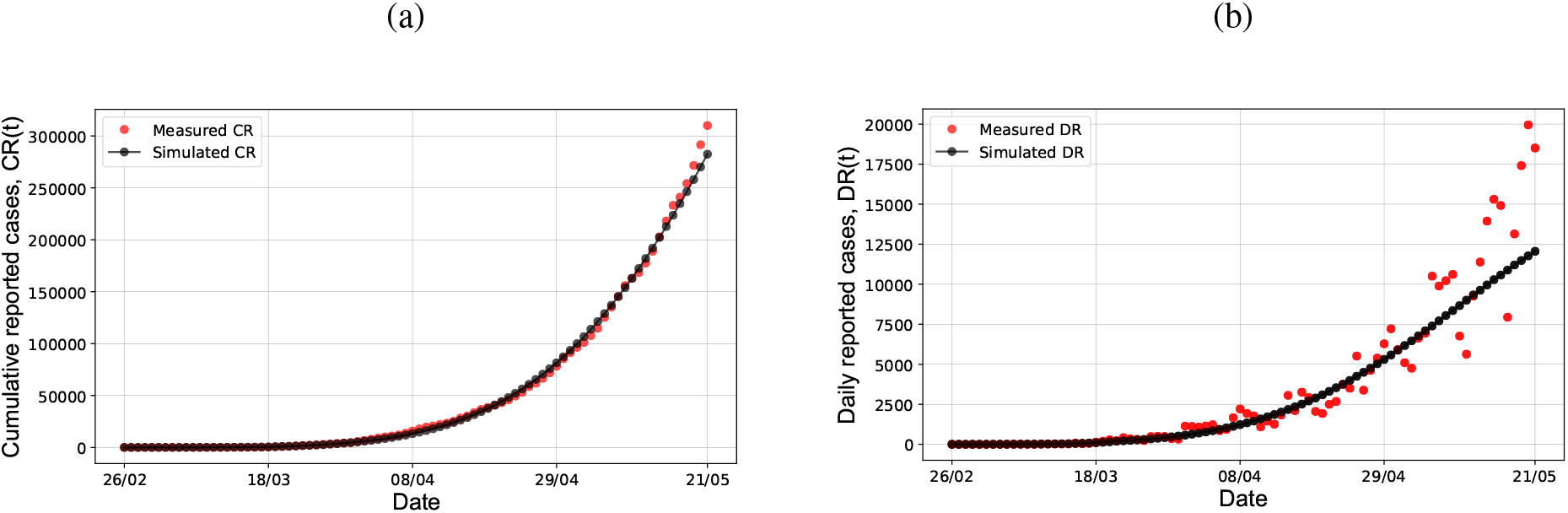
Monitoring results for cumulative (a) and daily (b) reported cases from PSO simulations, considering data from February, 26 2020 up to May 21, 2020 in Brazil, with *f*_0_ = 0.39123, *µ* = 0.0109 and *τ*_0_ = 1.426 · 10^−9^.

The result for the number of asymptomatic and symptomatic infectious cases given by the model states, *I*(*t*), *R*(*t*) and *U* (*t*), which are not measured variables, is also obtained (Figure 5). Infections may be transmitted from either asymptomatic individuals or unreported symptomatic individuals, which sum up more than 350,000 at this stage (May, 21 2020) of the pandemic in the country. It is known that most healthy people are likely to experience asymptomatic or mild cases; however, this fact makes the coronavirus spread control even harder.

**Figure 5:**
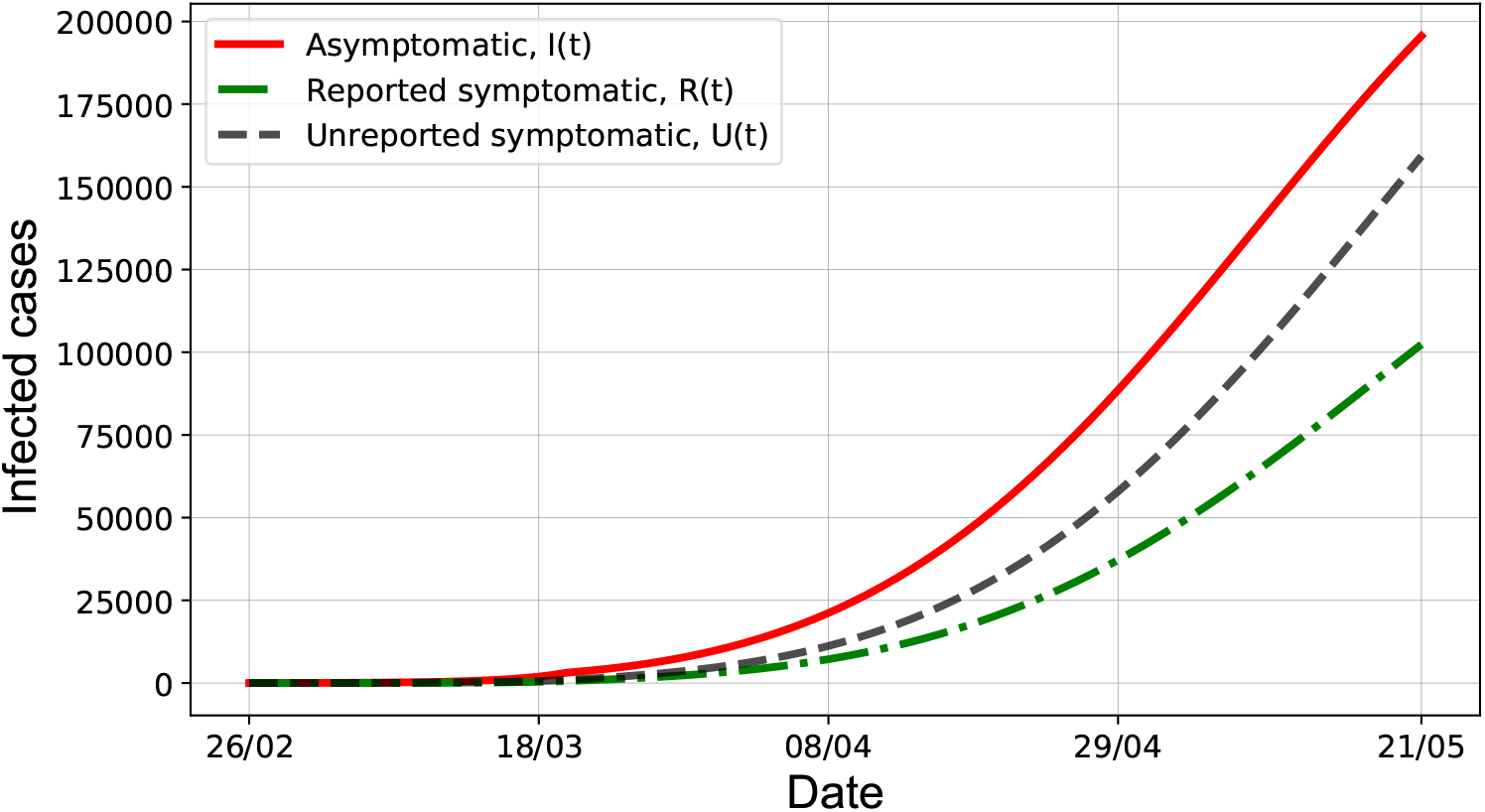
Monitoring results for reported and unreported cases from PSO simulations, considering data from February 26, 2020 up to May 21, 2020 in Brazil, for asymptomatic and symptomatic infectious cases, *I*(*t*), *R*(*t*) and *U* (*t*), with *f*_0_ = 0.39123, *µ* = 0.0109 and *τ*_0_ = 1.426 · 10^−9^.

Finally, in order to verify the predictive capability of this approach, we divided the problem into the learning and prediction steps as explained above. The best result of the simulations provided *f*_0_ = 0.39123, *µ* = 0.0109 and *τ*0 = 1.426 10^−9^. In Figure 6, from May 8, 2020, the prediction for 14 days is compared to the measured (or observed) values of the reported cases. We can see again that the SIRU model can be adjusted to represent the Brazil data set, since there is an excellent agreement with the data used in the learning step. However, the prediction step shows a poor performance when one desires to use the estimated parameters in the learning step. This is probably because any pandemics is a dynamic process depending on many factors, such as individual behaviour against isolation enforcement, application rate of test kits and processing of information. Since we used a PSO algorithm that does not consider time-varying parameters and uncertainties, this approach failed to exhibit a different trend which was not present in the training data.

**Figure 6:**
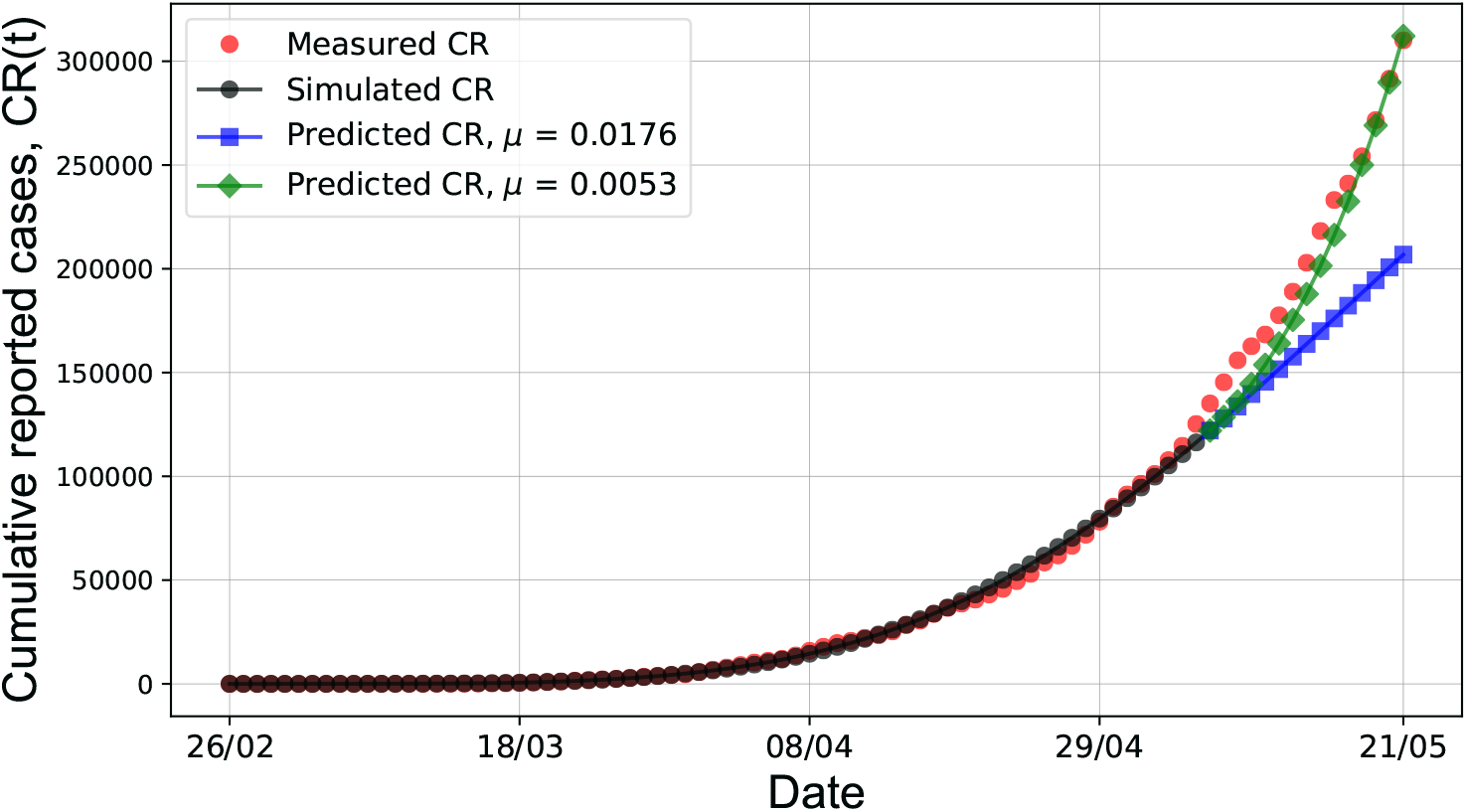
Simulation and prediction results from PSO simulations compared with the measured number of reported cases in Brazil from May 08, 2020 up to May 21, 2020. Different changes on were carried out to improve the performance of the prediction step.

The development of a reliable forecasting model is fundamental to allow public policies to be adopted in advance, in order to avoid further increasing the number of infected individuals. So, proposing a different scenario to predict properly may be interesting to show that the SIRU model can represent this pandemic. In this sense, we consider that the most uncertain parameter of the model would be the attenuation factor *µ*, which depends on the quarantine and isolation measures to decrease the infection rate. Keeping *f*0 constant at the estimated value, we checked for different changes on *µ* to find a scenario in which the performance of the prediction phase step could be improved. According to the result in Figure 6, we found that reducing substantially the attenuation factor by 70% would suffice to improve the prediction for the reported cases. Such result highlights the importance of considering state and parameter uncertainties. In this regard, we expect that the particle filter allows reaching a better prediction performance, what would make forecast analysis more reliable (or, at least, provide better understanding of the dynamic behaviour of the parameters)or, at least, understanding the time variation and range of the parameters. Anyway, the best result of the PSO algorithm is used merely here for comparison with the PF simulations, and to provide an initial estimate of the model parameters.

### 4.3 Estimation from PF simulations

We used the algorithm of Liu & West for the particle filter technique in the estimation problem. Such algorithm is suitable for the joint estimation of state variables and model parameters. The number of particles *N_part_* was set arbitrarily to 5000 and the results from the PSO simulations were considered as initial guesses for the parameters.

Gaussian prior probability densities were assumed for the parameters *f*0 and *µ* with standard deviations corresponding to approximately 1% of the means of each parameter. For the effect of the model uncertainty, we tuned throughout the simulations 0.001% for the number of individuals susceptible to infection, *S*(*t*), and 5% for the remaining states, *I*(*t*), *R*(*t*) and *U* (*t*) – such percentage is related to the previous value of the respective state. Besides that, the uncertainty level for the observed cumulative cases, *CR*(*t*), changes over time. It started at *σ_CR_* = 30% in relation to the measured value and, as of March 17, 2020 when isolation was enforced, it was decreased in the simulation by a step-change to *σCR* = 3%. To present the results, we calculated the estimation error, 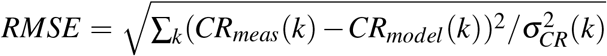, and 95% credibility level for each state variable and parameter was calculated as the 2.5% and 97.5% quantiles.

The posterior distributions of each state and parameters estimated on May 07, 2020 (last day of the learning step) provided the respective initial condition for the prediction step. Lastly, we also performed an additional step to forecast new Covid-19 cases during 120 days (from May 22, 2020 to September 17, 2020). In the prediction and forecast horizons, the model parameters are updated via random walk model.

Using the full data set (February 26, 2020 to May 21, 2020), the estimation with the algorithm of Liu & West significantly improved the agreement of the simulated values with the measured values (Figure 7), especially for those measured points near the end of the considered period. This was possible due to the dynamic estimation of the parameters (Figure 8 a and b), which were varied over time from the initial guess. The attenuation factor was increased about 30% from the initial guess, showing that isolation has been effective at some extent decreasing the transmission rate, *τ*(*t*). The fraction of report shows that over 60% of symptomatic infectious cases were not reported in this period. The improvement obtained in comparison to the PSO result can be seen from the estimation error *RMSE* (Figure 8d), which was consistently decreased. Regarding the infectious cases, the estimation is alarming since the group of potential transmitters is increasing (Figure 9). The number of the infectious population that may not have been diagnosed (that is, asymptomatic and unreported symptomatic individuals) is estimated to be more than 450,000 cases. Hence, it is worthy to point out that massive, rapid tests and restrictive measures are paramount to curb the spread of the virus.

**Figure 7:**
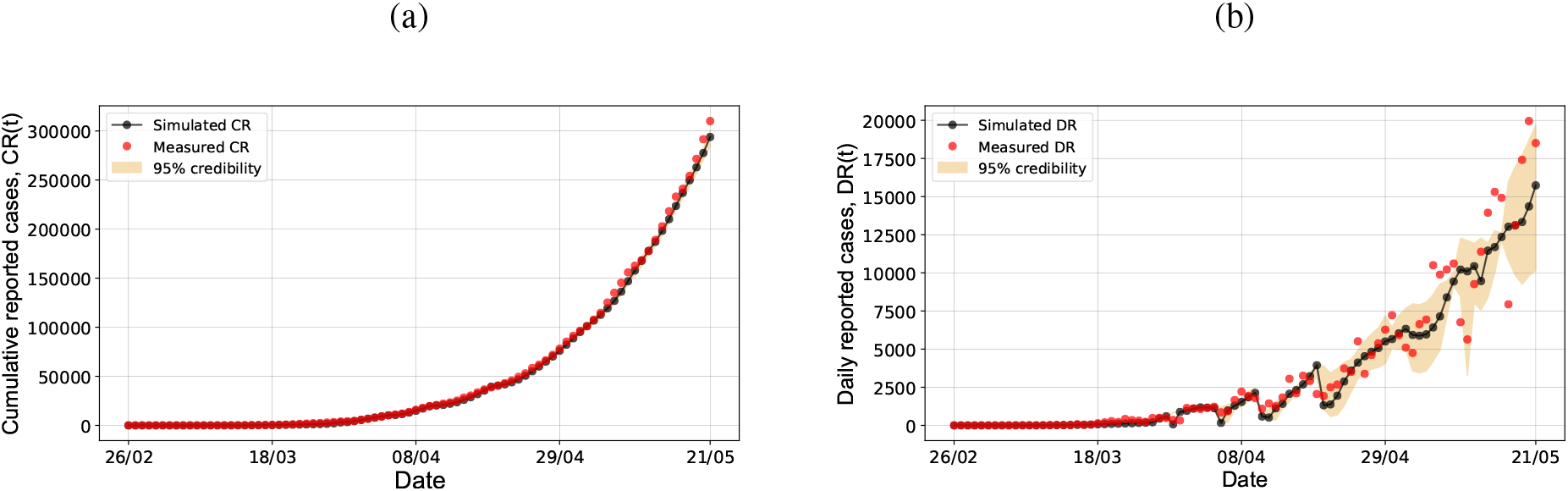
Monitoring results for (a) cumulative cases and (b) daily reported cases from the PF simulations, considering data from February 26, 2020 up to May 21, 2020 in Brazil.

**Figure 8:**
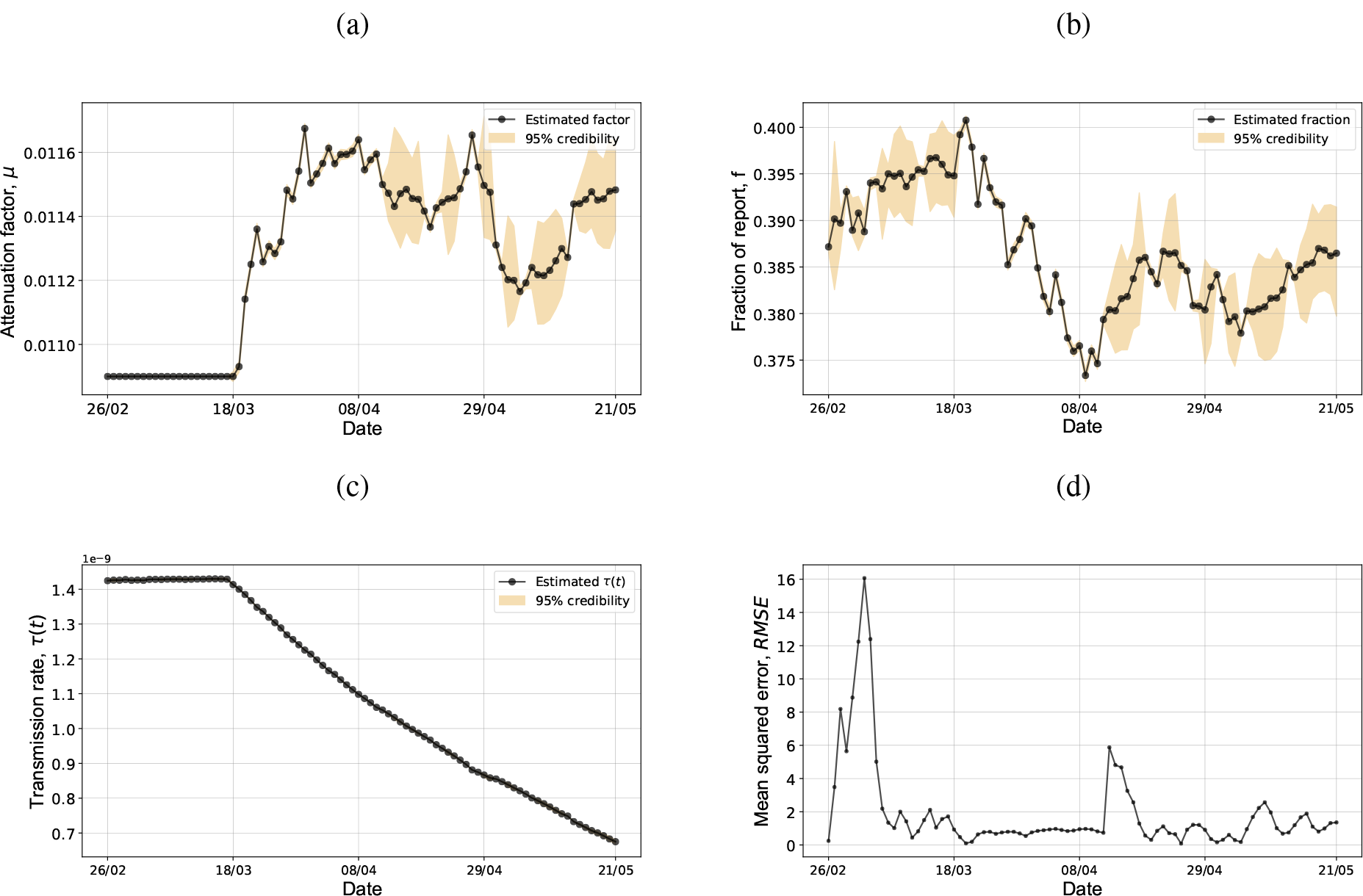
Dynamic results of the SIRU model parameters: (a) attenuation factor, *µ*; (b) fraction of report, *f*(*t*) = *f*_0_; (c) transmission rate, *τ*(*t*) = *τ*_0_*exp*[−*µ*(*t* − *N*)]; and (d) the mean squared error, RMSE, from the PF simulations, considering data from February 26, 2020 up to May 21, 2020 in Brazil.

**Figure 9:**
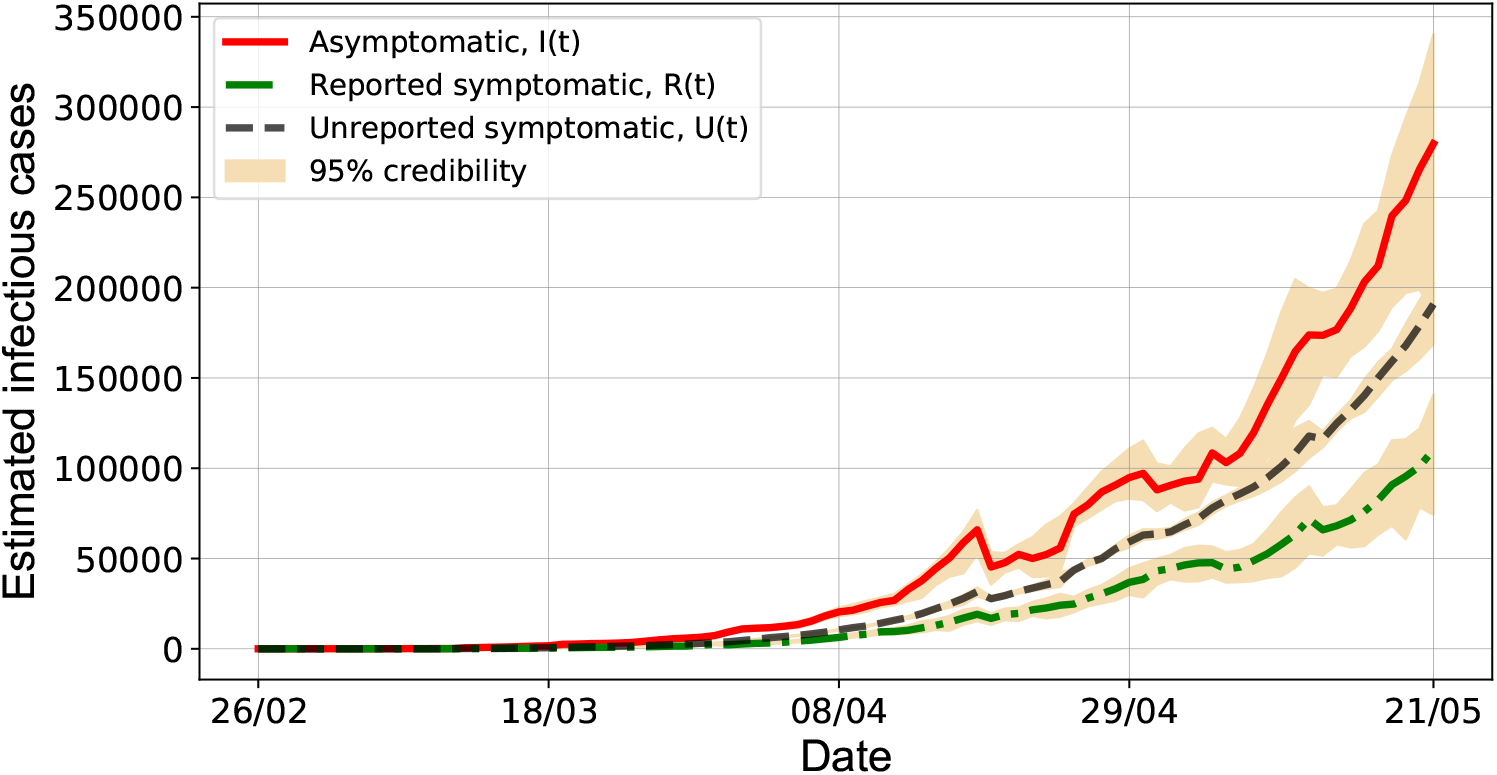
Monitoring results for reported and unreported cases from the PF simulations, considering data from February 26, 2020 up to May 21, 2020 in Brazil, for asymptomatic and symptomatic infectious cases, *I*(*t*), *R*(*t*) and *U* (*t*).

The capability of the model to predict was analyzed using the available data of reported cases in Brazil from February 26, 2020 up to May 7, 2020. According to Figure 10, the adjustment of the parameters found in the learning step is suitable to predict accurately the pandemic evolution up to May 18, 2020 (10 days ahead), which would have presented in advance the notification of more than 115,000 cases.

**Figure 10:**
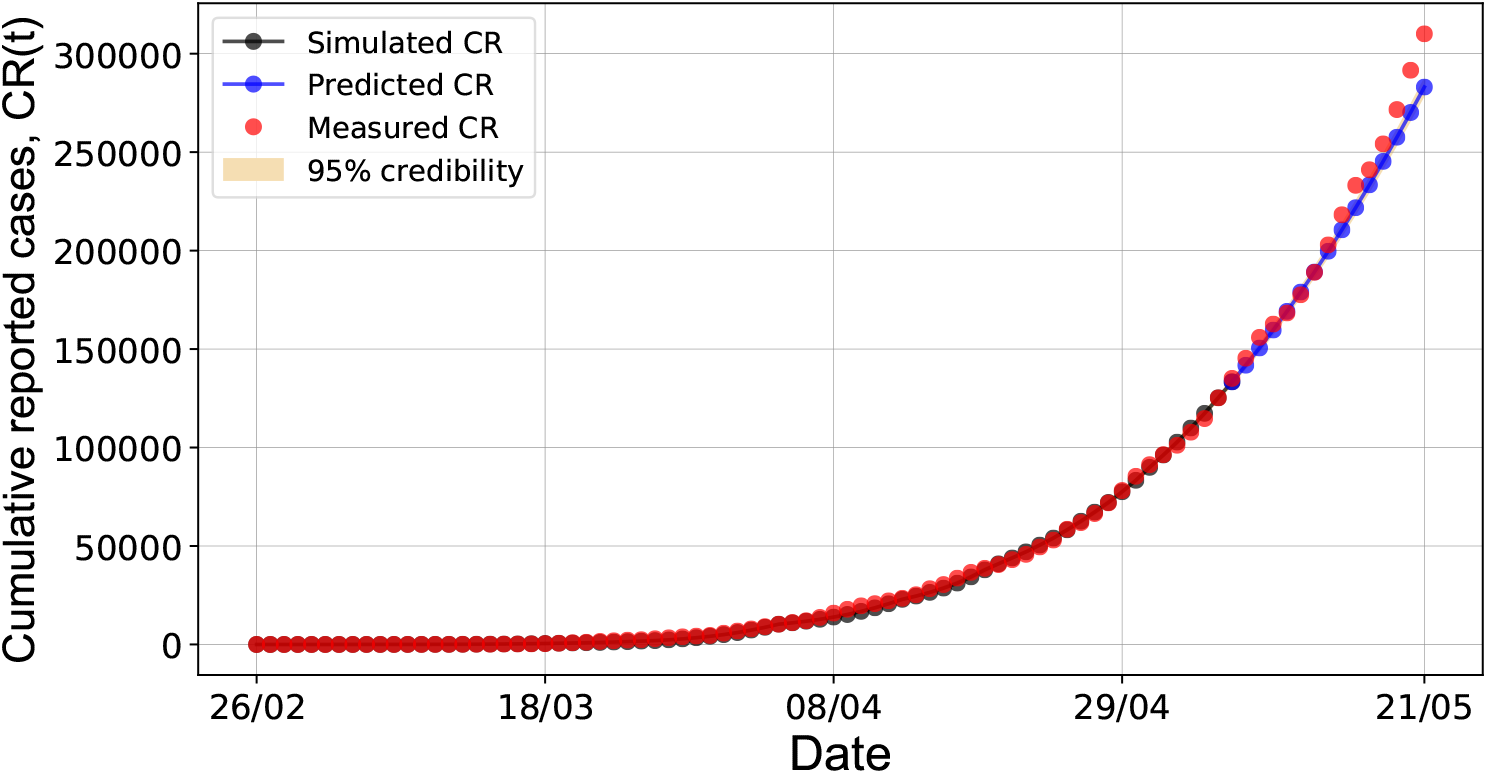
Validation of the predictive capability of the PF simulations for the measured number of reported cases in Brazil, considering data from May 08, 2020 up to May 21, 2020.

From this result, we simulated the model considering 120 days from May 21, 2020 to forecast new cases. The dynamic behaviour of the parameters in both steps can be seen in Figure 11. The forecasted values for the daily reported cases and for the infectious cases are shown in Figures 12 and 13, in which we highlighted each analysis step (estimation, prediction and forecast). The result suggests that the epidemic peak in Brazil are due to be reached around mid-June, 2020, with almost 25,000 cases being notified a day. Specifically, it can inferred that the number of asymptomatic individuals may increase up to 400,000 cases and, more importantly, the number of unreported symptomatic cases around 460,000 may be twice greater than the number of reported symptomatic. In brief, this means that number of people requiring medical assistance will increase dramatically in the future. According to the forecast, the pandemics in Brazil is expected to slow down beyond the peak of daily number of cases and the number of cumulative reported cases to be more than 1,500,000 (Figure 14). So, if such numbers impose a burden on the health system, it is important to enhance isolation measures, as quarantine and social distancing. As Brazil is a very large country, the spread of the virus is supposed to vary from region to region, such way that community containment measures may be established locally at different moments.

**Figure 11:**
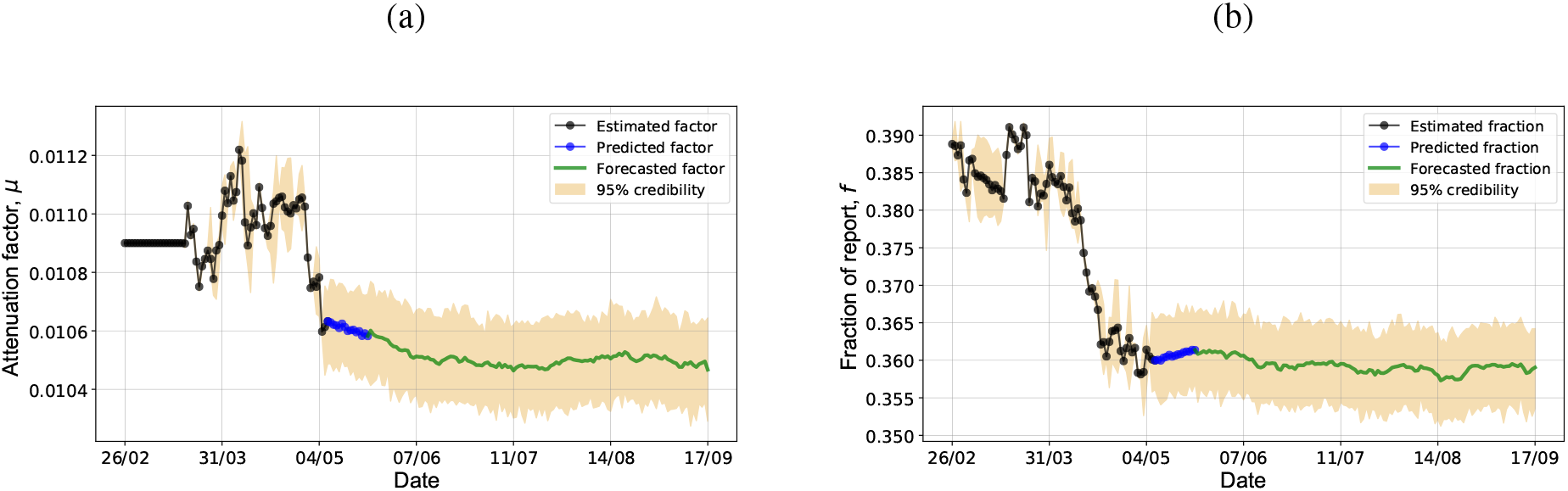
Dynamic behaviour of the parameters during the prediction and forecasting steps: (a) attenuation factor, *µ*; (b) fraction of report, *f* (*t*) = *f*_0_.

**Figure 12:**
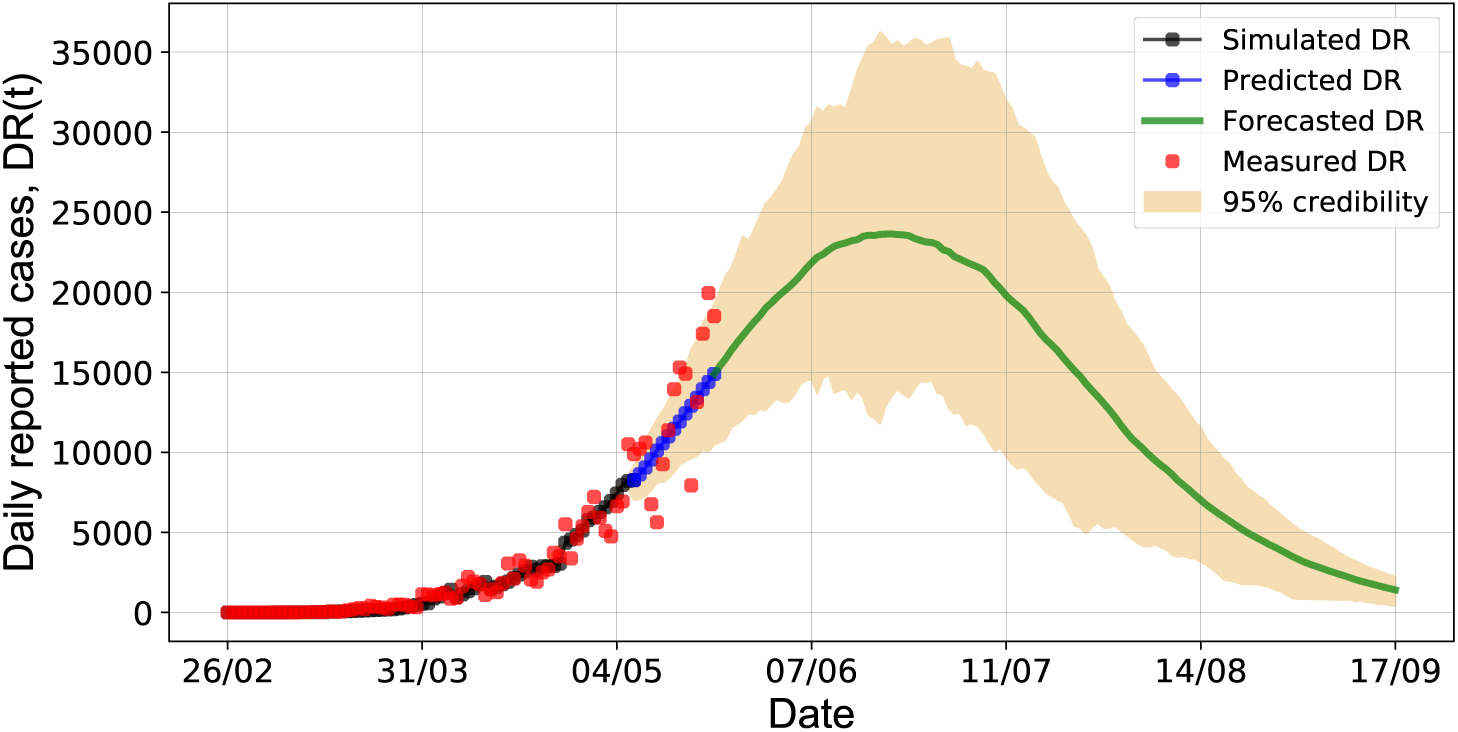
Forecast result for the daily reported cases in Brazil up to September 17, 2020.

**Figure 13:**
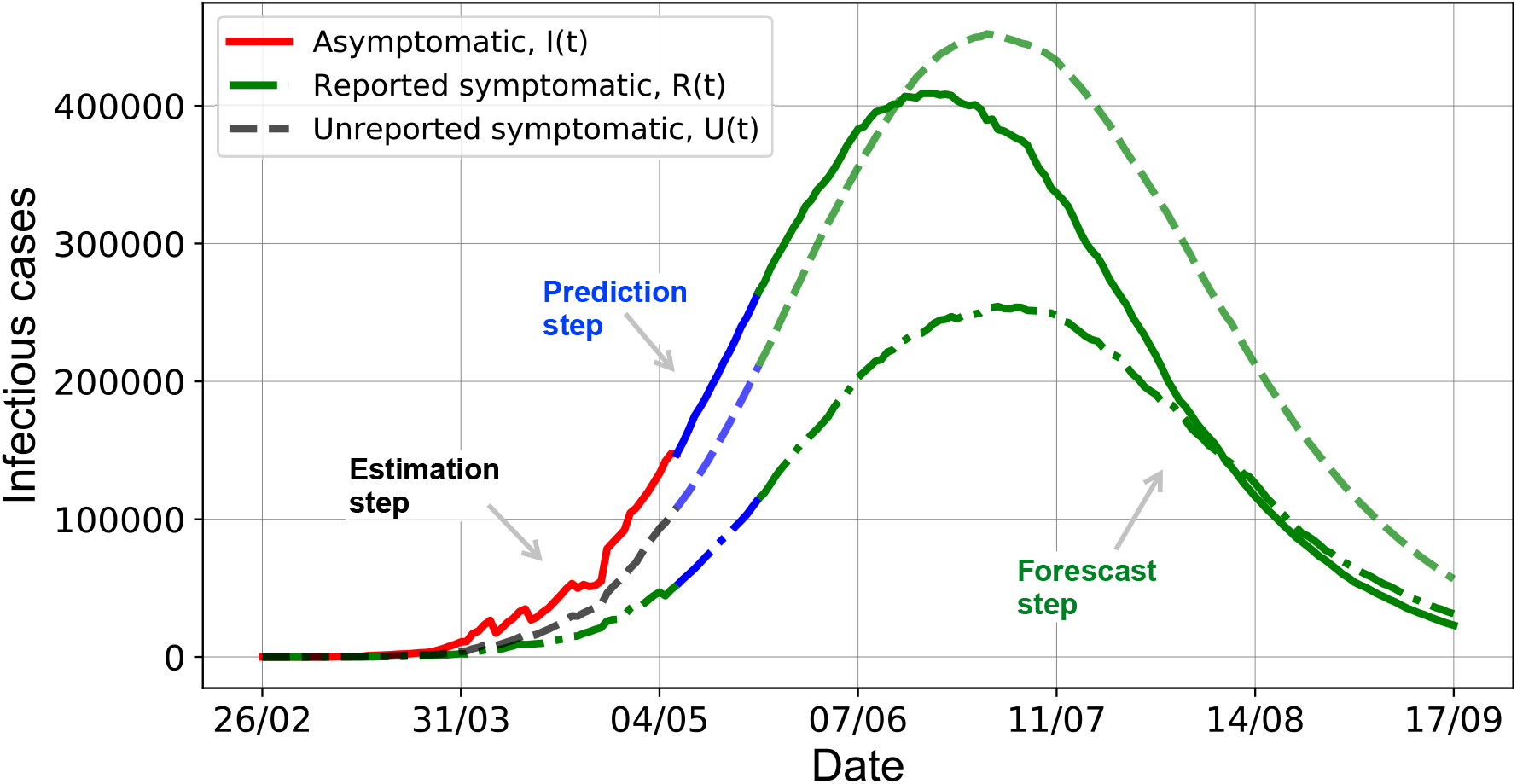
Forecast result for the number of infectious cases up to September 17, 2020.

**Figure 14:**
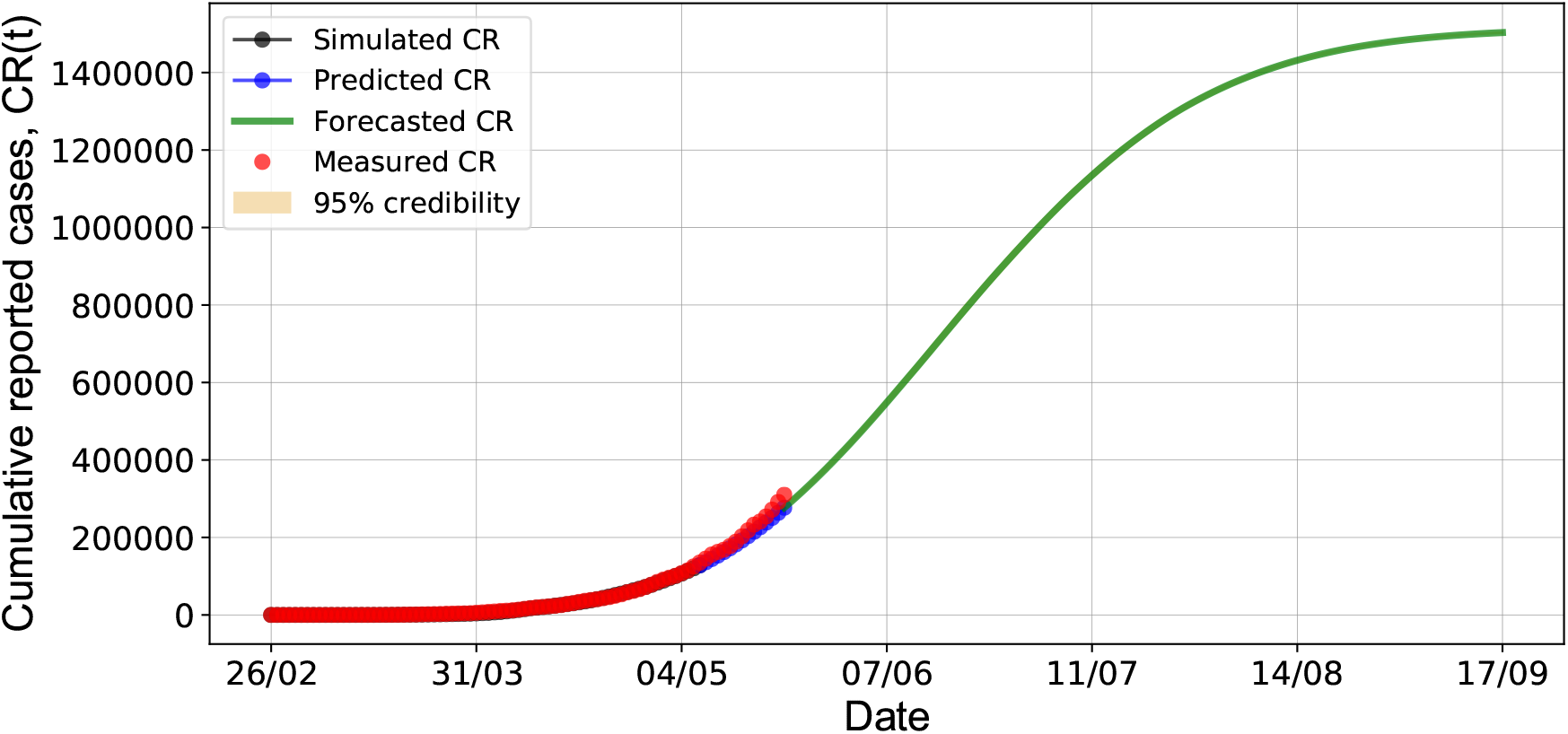
Forecast result for the cumulative reported cases up to September 17, 2020.

## 5 Conclusions

Since the coronavirus outbreak has been evolving rapidly, this work considered a numerical application to predict the evolution of a Covid-19 epidemic in Brazil. We implemented the model SIRU to represent the coronavirus spread and used the cumulative reported cases as measurement. For the estimation problem, an inverse problem was solved under the Bayesian framework with particle filter. Specifically, we used the algorithm of Liu & West to perform a joint estimation of states and parameters. Besides that, particle swarm optimization based simulations provided initial guesses for this problem. The model was calibrated to represent the number of reported cases and validated using two-week real data. Afterwards, we performed a forecast step to predict new cases of the disease. It was found that the pandemic peak is expected to take place in mid-June, 2020 with about 25,000 news cases a day and over 1,500,000 individual will have tested positive for Covid-19 up to September, 2020. Such result reinforces the need for social distancing measures and for diagnostic tests to decrease the transmission rate and the number of infected people in severe condition.

## Data Availability

All the data used in this manuscript were obtained from the Brazilian Ministry of Health available at its webpage.

https://covid.saude.gov.br/

## Notes

### Competing Interest Statement

The authors have declared no competing interest.

### Funding Statement

No external funding was received.

### Author Declarations

This manuscript does not involve research involving human subjects. We considered a numerical application to predict the evolution of a Covid-19 epidemic in Brazil, using a SIRU-type model to represent the coronavirus spread and a Bayesian framework to perform a joint estimation of states and parameters.

